# Can prognostic factors for indirect muscle injuries in elite football (soccer) players be identified using data from preseason screening? An exploratory analysis using routinely-collected periodic health examination records

**DOI:** 10.1101/2021.01.15.21249653

**Authors:** Tom Hughes, Richard D. Riley, Michael J. Callaghan, Jamie C. Sergeant

## Abstract

This study used periodic health examination (PHE) data from 134 outfield elite male football players, over 5 seasons (1^st^ July 2013 to 19^th^ May 2018). Univariable and multivariable logistic regression models were used to determine prognostic associations between 36 variables and time-loss, lower extremity index indirect muscle injuries (I-IMIs). Non-linear associations were explored using fractional polynomials. During 317 participant-seasons, 138 I-IMIs were recorded. Univariable associations were determined for previous calf indirect muscle injury (IMI) frequency (OR=1.80, 95% confidence interval (CI) = 1.09 to 2.97), hamstring IMI frequency (OR=1.56, 95% CI=1.17 to 2.09), if the most recent hamstring IMI occurred > 12 months but < 3 years prior to PHE (OR= 2.95, 95% CI = 1.51 to 5.73) and age (OR =1.12 per 1-year increase, 95% CI = 1.06 to 1.18). Multivariable analyses demonstrated that if a player’s most recent previous hamstring IMI was >12 months but <3 years prior to PHE (OR= 2.24, 95% CI = 1.11 to 4.53), then this was the only variable with added prognostic value over and above age (OR=1.12 per 1-year increase, 95%CI = 1.05 to 1.18). Allowing non-linear associations conferred no advantage over linear ones. Therefore, PHE has limited use for injury risk prediction.

## INTRODUCTION

Periodic health examination (PHE), or screening, is a well-established clinical evaluation strategy in elite football.^1^ Typically during PHE, players undertake various medical, musculoskeletal, functional and performance tests^2^ during preseason and in-season periods.^1^ PHE allows opportunities for general health surveillance, identification of salient pathology^3^ and monitoring of rehabilitation or performance.^4^ PHE is also assumed to have an important role in the development of injury prevention strategies.^5 6^ In particular, although PHE cannot establish specific causal factors for injuries,^4^ it is perceived to be useful for the prediction of future injury risk in athletes,^2 4^ where accurate predictions could help to identify individuals who may require interventions that are designed to reduce risk.^7^

To make such predictions, prognostic factors are required which, in the context of football, could be any variables, characteristics or measurements available at or derived from PHE (e.g. medical history, leg strength or range of motion tests) that are associated with increased injury risk.^4^ The predictive power of a single prognostic factor is usually limited on its own.^8^ 9 However, if several prognostic factors are used in combination within a multivariable prognostic model, it may be possible to produce useful individualised risk estimates.^8 10^

Because the predictive function of PHE remains unsubstantiated^3 11^ and given that indirect (non-contact) muscle injuries (IMIs) are a significant problem observed in elite football (accounting for 30.3% to 47.9% of all injuries),^12-16^ we have recently developed and internally validated a multivariable prognostic model to predict individualised lower extremity IMI risk in elite players using PHE data.^17^ However, sample size limitations meant that only 10 candidate prognostic factors could be considered in our model and these were selected using data quality assessment, clinical reasoning, or on the basis of our related systematic review.^11^ The performance of our model was modest and we concluded that its use would not be beneficial in clinical practice.^17^

Furthermore, our previous systematic review also highlighted several methodological limitations of the current evidence, which specifically included inadequate reporting of outcomes, prognostic factor measurement and reliability.^11^ Additionally, while most studies performed appropriate statistical analyses, continuous prognostic factor measurements were often categorised^18-21^ and non-linear associations were not investigated,^18-24^ which does not conform to current methodological recommendations.^25-27^

To assist the development of future prognostic models, there is a clear need to ascertain the existence of robust and novel prognostic factors that have an association with IMIs.^17^ Therefore, using routinely collected data from a 5-season period, we conducted an exploratory cohort study to examine: 1) prognostic associations between PHE-derived variables and IMI outcomes in elite footballers, using a wider set of candidates than had previously been considered^17^ and; 2) the prognostic value of these PHE-derived variables over and above standard anthropometric variables of age (which has previously confirmed prognostic value^11 17^), height and weight. In particular, both linear and non-linear associations between candidate variables and outcomes were explored which, to the best of our knowledge, has not been conducted previously.

## METHODS

The methodology has been described in a published protocol^28^ so will only be briefly outlined. This study was registered on ClinicalTrials.gov (NCT03782389) and was reported according to the Reporting Recommendations for Marker Prognostic Studies (REMARK).^29^ Given the number of PHE-related variables examined, our study should be viewed as exploratory, but we emphasise that this an important phase in prognostic factor research.^9 30^

### Data Sources

This study was of retrospective cohort design. Eligible participants were identified from a population of male elite footballers, aged 16-40 years old at Manchester United Football Club. A database was created using routinely collected injury records and preseason PHE data over 5 seasons (1^st^ July 2013 to 19^th^ May 2018). For each season (which started on 1^st^ July), participants completed a mandatory PHE during the first week of the season and were followed up to the last first team game of the season.

The PHE process typically included: 1) anthropometric measurements; 2) a review of medical and previous injury history; 3) musculoskeletal examination tests; 4) functional movement and balance tests; 5) strength and power tests. The PHE test order was self-selected by each player and a standardised warm up was not implemented, although players could undertake their own warm up procedures if they wished. Each component of PHE was standardised according to a written protocol and was examined by physiotherapists, sports scientists or club medical doctors. The same examiners performed the same test every season to eliminate inter-tester variability. No examiner attrition occurred throughout the data collection period. If a participant was injured at the scheduled time of PHE, a risk assessment was completed by medical staff and participants only completed tests that were deemed appropriate and safe for the participant’s condition; examiners were therefore not blinded to injury status.

### Eligibility criteria

During any season, participants were eligible if they: 1) were not a goalkeeper; 2) participated in PHE for the relevant season. Participants were excluded if they were not under contract to the Club at the time of PHE.

### Ethics and Data Use

Informed consent was not required as data were captured from the mandatory PHE completed through the participants’ employment. The data usage was approved by the Club and the Research Ethics Service at the University of Manchester.

### Participant involvement

Participants were not involved in the study design.

### Outcome

The outcome was any time-loss, index lower extremity IMI (I-IMI) sustained by a participant during match play or training, which affected any lower abdominal, hip, thigh, calf or foot muscle groups and prohibited future match or training participation.^31^ I-IMIs were confirmed and graded by a club doctor or physiotherapist according to the previously validated Munich Consensus Statement for the Classification of Muscle Injuries in Sport,^32 33^ during routine assessments undertaken within 24h of injury occurrence. The medical professionals were not blinded to PHE data at diagnosis.

### Sample size

Our sample size of 317 participant-seasons (with 138 I-IMI events) had 80% power to detect an adjusted odds ratio (OR) of at least 1.6 for a 1 standard deviation increase in a variable of interest, conservatively assuming a correlation of 0.5 with the adjustment variables of age, height and weight (see supplementary file 1 for the sample size calculation).^34^

### PHE-derived Candidate Variables

The dataset contained 60 variables^28^ that were eligible for analysis unless there were >15% missing observations or if reliability (where applicable) was reported as fair to poor (that is, Intraclass Correlation Coefficient (ICC) < 0.70).^28 35^ If any variables did not meet these eligibility criteria, they were excluded (supplementary file 2). Collinearity between eligible variables was assessed with a scatterplot matrix; this was evident when tests were used to measure right and left limbs independently.^28^ In these circumstances, composite variables were created for between-limb differences and the mean of the test measurements for both limbs, as described in the study protocol.^28^

Of the remaining eligible variables, 10 were used in our multivariable prognostic model study (represented by 12 parameters).^17^ With the exception of age at PHE (which was used for adjustment purposes in this study), these candidates were therefore excluded.^28^ The final number of candidate variables included for exploratory analysis was 36. Table 1 summarises all included variables with their measurement units and data type, as well as the methods and reliability of measurement.

**Table 1:**
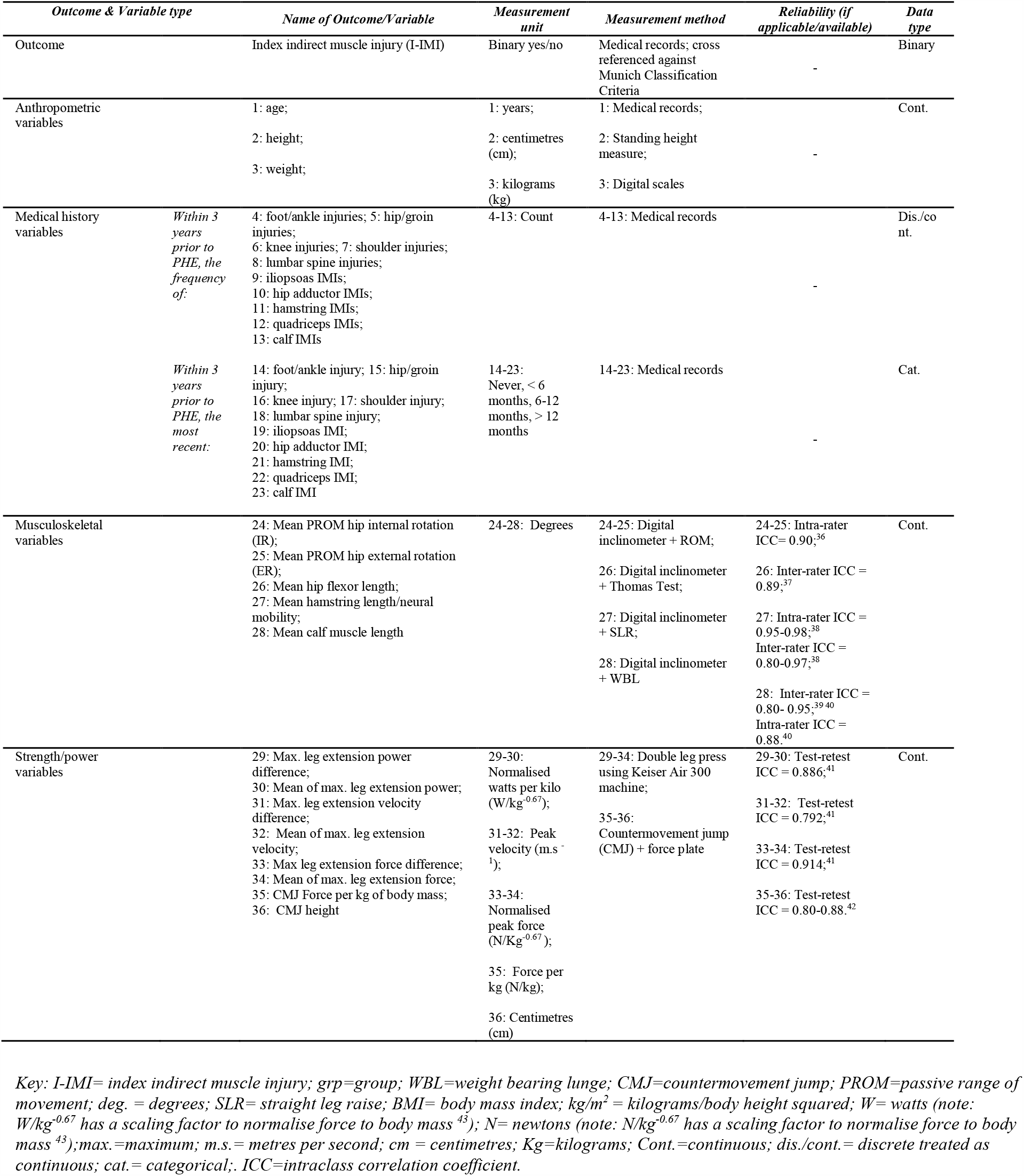
List of candidate variables with a summary of the units of measurement, methods and reliability of measurement and data type.

### Statistical analysis

#### Data handling – outcome measures

Each participant-season was treated as independent. If an I-IMI occurred, the participant’s outcome was determined for that season and they were no longer considered at risk. In these circumstances, participants were included for further analysis at the start of the consecutive season, if still eligible. Any upper limb IMI, trunk IMI or non-IMI injuries were ignored and participants were still considered at risk.

Eligible participants who were loaned to another club throughout that season, but had not sustained an I-IMI prior to the loan were still considered at risk. I-IMIs that occurred whilst on loan were included for analysis. Permanently transferred participants (who had not sustained an I-IMI prior to the transfer), were recorded as not having an I-IMI during the relevant season and exited the cohort at the season end.

#### Data Handling – Missing data

Missing values were assumed to be missing at random (i.e. missingness could be predicted conditional on other known variables).^28^ The continuous parameters generally demonstrated non-normal distributions, so were transformed using normal scores^44^ to approximate normality before imputation, and back-transformed following imputation.^45^ Multivariate normal multiple imputation was performed, using a model that included all candidate variables and I-IMI outcomes. Fifty imputed datasets were created in Stata 15.1 (StataCorp LLC, Texas, USA) using the ‘*mi impute*’ command.

#### Univariable and multivariable analyses

All data were analysed in the form that they were recorded. In particular, variables that were recorded as continuous were kept continuous and not categorised, to avoid a loss of prognostic information.^25^ Univariable logistic regression models were used to estimate the unadjusted linear associations between I-IMIs and each candidate variable. Multivariable logistic regression models were also used to estimate the linear association between I-IMIs and each variable, after adjustment for age (which has confirmed prognostic importance^11 17^), height and weight (which were both considered as potential confounders for I-IMIs and PHE-derived candidates). All parameter estimates were averaged across all imputed datasets using Rubin’s Rules^46^ and were computed using the ‘*mim*’ module in Stata 15.1. Statistical significance thresholds were used to indicate the strength of exploratory evidence against null associations, where p-values of : 1) <0.05 indicated strong evidence and the factor was considered significant; 2) 0.05 to 0.10 indicated weak evidence and; 3) >0.10 indicated little or no evidence.^47^ Prognostic importance was also considered by checking the magnitude of prognostic effects encompassed by the width of 95% confidence intervals.

For all variables, non-linear associations with the outcome were also explored using fractional polynomials for the univariable and multivariable models; the fit of first and second order fractional polynomial models were evaluated against the fit of the standard logistic regression models.^48^ The parameter estimates were combined across all imputed datasets^49^ using Rubins Rules,^46^ with the automated ‘*mfpmi*’ algorithm in Stata 15.1, using a significance threshold set at p< 0.05. All analyses are summarised in Table 2.

**Table 2:** Summary of all statistical analyses performed

| A) Statistical analyses of I-IMI outcomes |  |  |  |  |  |
| --- | --- | --- | --- | --- | --- |
| *Analysis performed* | *Participant-seasons* | *Number of I-IMIs* | *Variables considered* | *Adjusted for:* | *Results* |
| Primary analysis, imputed data |  |  |  |  |  |
| A1: Univariable Logistic Regression/FPs | 317 | 138 | Individual models for I-IMI + each variable (1-36) | None | Table 4 |
| A2: Multivariable Logistic Regression/FPs | 317 | 138 | Individual models for I-IMI + each variable (4-36) | Variables 1-3 (age, height, weight) | Table 4 |
| Primary analysis, complete case data |  |  |  |  |  |
| B1: Univariable Logistic Regression/FPs | 265 | 115 | Individual models for I-IMI + each variable (1-36) | None | SF 5 |
| B2: Multivariable Logistic Regression/FPs | 265 | 115 | Individual models for I-IMI + each variable (4-36) | Variables 1-3 (age, height, weight) | SF 5 |
| Sensitivity analysis, imputed data |  |  |  |  |  |
| C1: Univariable Logistic Regression/FPs | 260 | 129 | Individual models for I-IMI + each variable (1-36) | None | SF 6 |
| C2: Multivariable Logistic Regression/FPs | 260 | 129 | Individual models for I-IMI + each variable (4-36) | Variables 1-3 (age, height, weight) | SF 6 |
| Sensitivity analysis, complete case data |  |  |  |  |  |
| D1: Univariable Logistic Regression/FPs | 217 | 106 | Individual models for I-IMI + each variable (1-36) | None | SF 7 |
| D2: Multivariable Logistic Regression/FPs | 217 | 106 | Individual models for I-IMI + each variable (4-36) | Variables 1-3 (age, height, weight) | SF 7 |

Key: I-IMI= index indirect muscle injury; FP= fractional polynomials; SF= supplementary file

#### Primary and sensitivity analyses

To determine the effect of imputation and player transfers on variable associations, the analyses were repeated: 1) as complete cases analyses; and 2) as sensitivity analyses excluding participant-seasons for participants who were loaned or transferred (performed as both multiple imputation and complete case analyses). All primary complete case and sensitivity analyses are also summarised in Table 2.

## RESULTS

### Participants

During the five seasons, 134 participants were included, contributing 317 participant-seasons and 138 IMIs in the primary analysis (Figure 1). Three players were classified as injured at the time of PHE (which affected three participant-seasons). This meant they were unavailable for selection for training or matches at that time. However, these players had commenced football specific, field-based rehabilitation so also had similar exposure to training activities to uninjured players and were therefore included in the cohort because it was reasonable to assume that they could also be considered at risk of an I-IMI event.

**Figure 1:**
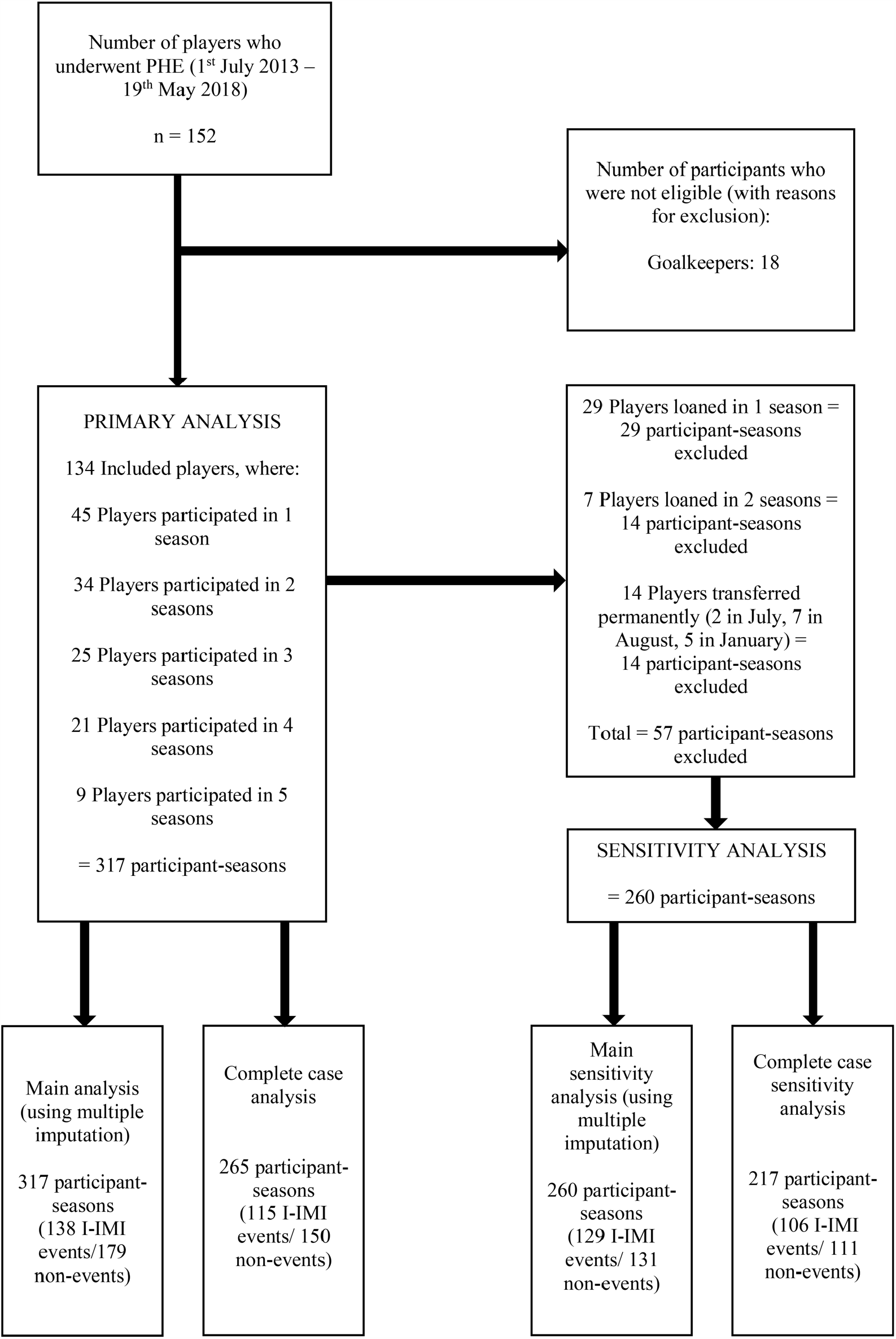
Participant flow chart *Key: n=number of participants; I-IMI=index indirect muscle injury*

For the sensitivity analyses (excluding loans and transfers), 260 independent participant-seasons with 129 IMIs were included; 36 participants were transferred on loan, while 14 participants were permanently transferred during a season, which excluded 57 participant-seasons (Figure 1).

Table 3 summarises the participant characteristics and candidate variable values for participants included in the primary analyses. All values were similar to those included in the sensitivity analyses (supplementary file 3).

### Missing data and multiple imputation

Data were complete for age and all past medical history variables (Table 3). For all other candidates, the proportion of missing data ranged from 5.68% (for height and weight) to 14.20% (for the mean and between limb differences of maximal leg extension power and force) (Table 3). For all continuous variables, the distribution of imputed values approximated the observed values (supplementary file 4), therefore confirming their plausibility.

**Table 3.**
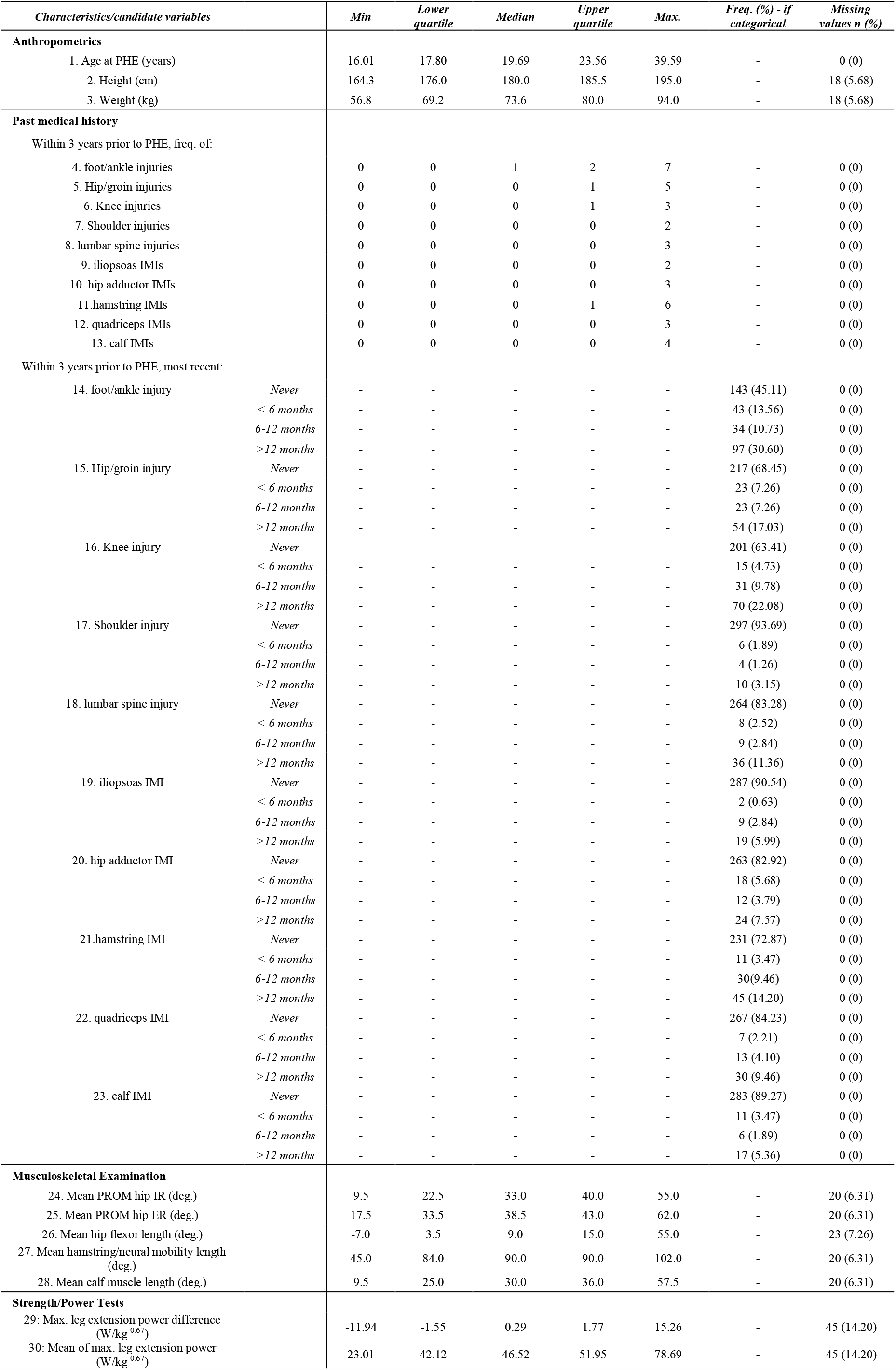
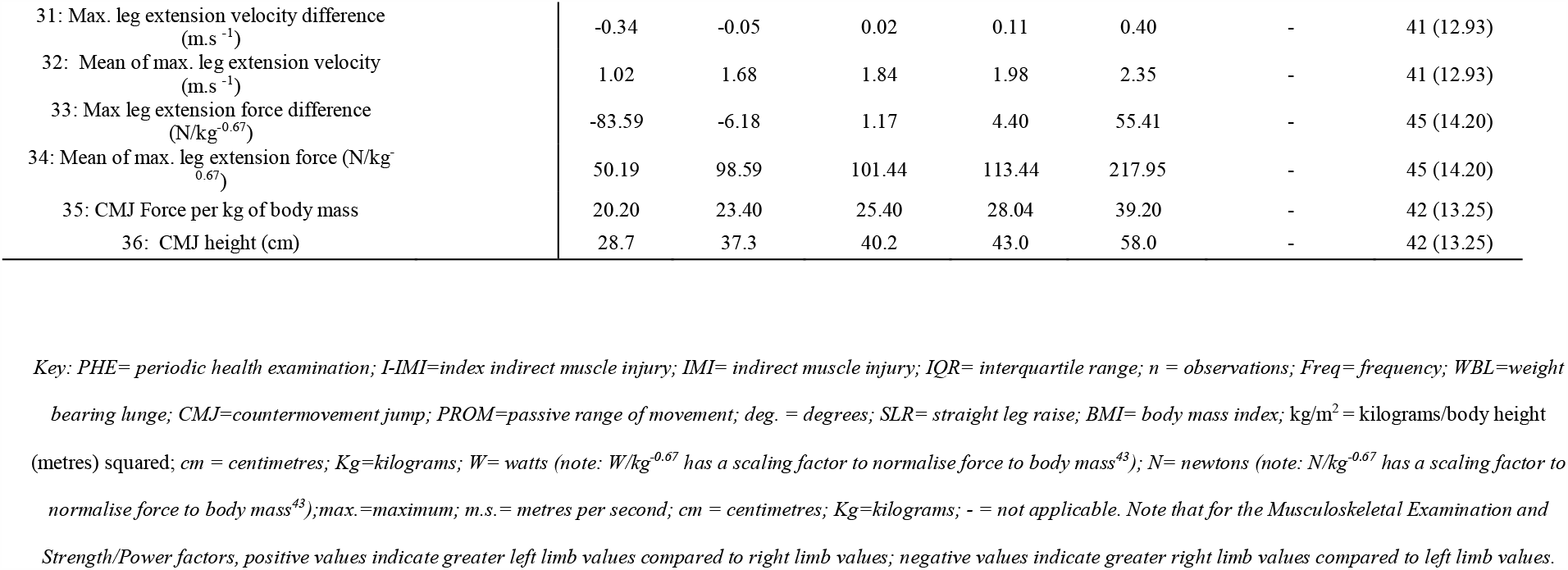
Characteristics of included participants

### Univariable analyses

Table 4 shows the results of the univariable analyses. The continuous variables of age (OR= 1.12 for a 1-year increase, 95% confidence interval (CI)=1.06 to 1.18, p<0.001), weight (OR=1.03 for a 1kg increase, 95% CI=1.00 to 1.07, p=0.03) and mean hip IR PROM (OR=0.97 for a 1-degree increase, 95% CI = 0.95 to 0.99, p=0.01) showed a significant but modest association with I-IMIs. The narrow CIs indicated that these estimates were relatively precise. Linear associations were the best fit for all these continuous variables. Significant associations with larger OR estimates were observed for previous calf IMI frequency (OR=1.80, 95% CI = 1.09 to 2.97, p=0.02), hamstring IMI frequency (OR=1.56, 95% CI=1.17 to 2.09, p<0.001), and if the most recent hamstring IMI occurred more than 12 months but less than 3 years prior to PHE (OR= 2.95, 95% CI = 1.51 to 5.73, p<0.001). The wider CIs for these estimates indicated greater imprecision about the prognostic effect; this may because these candidates were either discrete or categorical, rather than continuous.

**Table 4:**
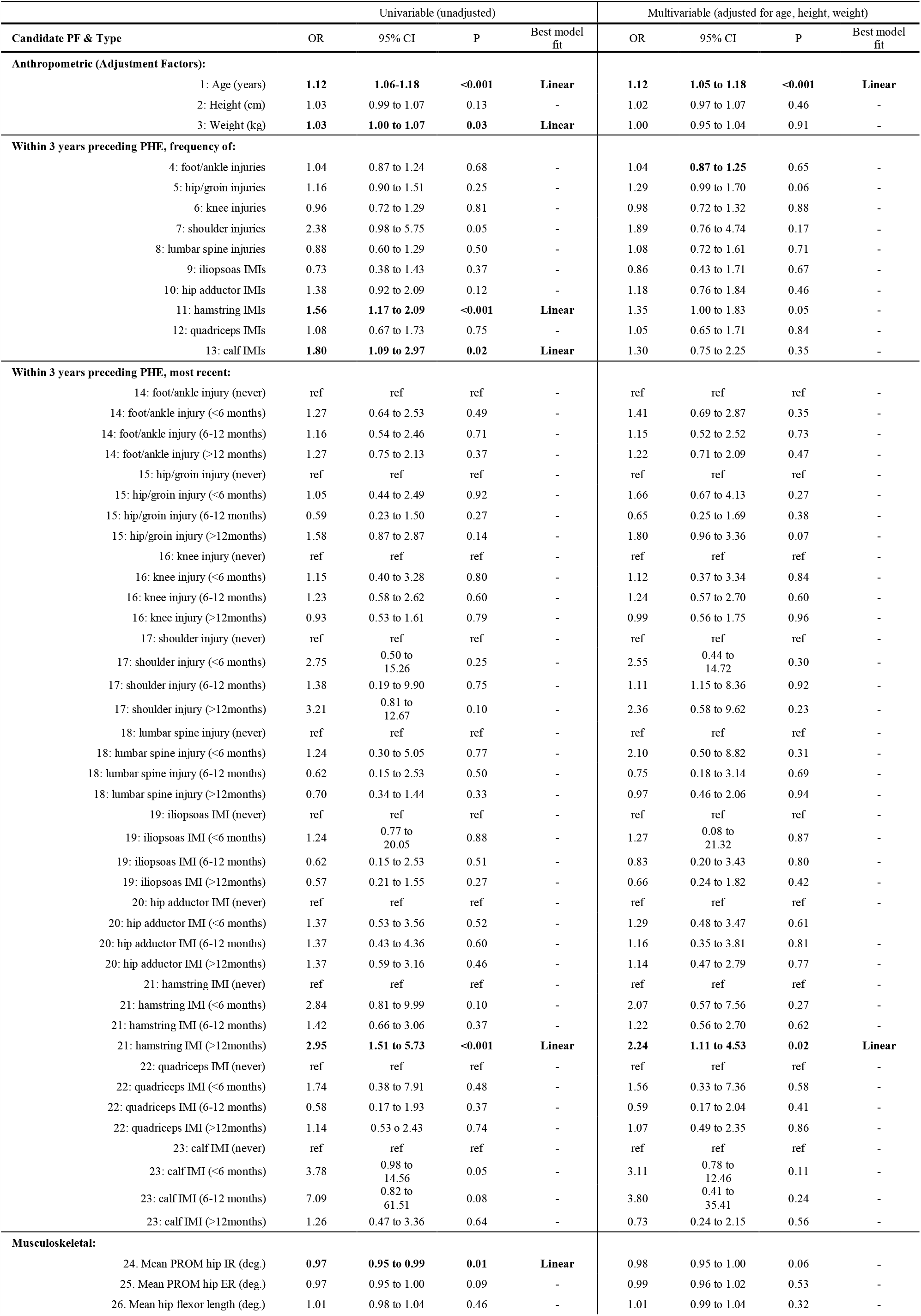
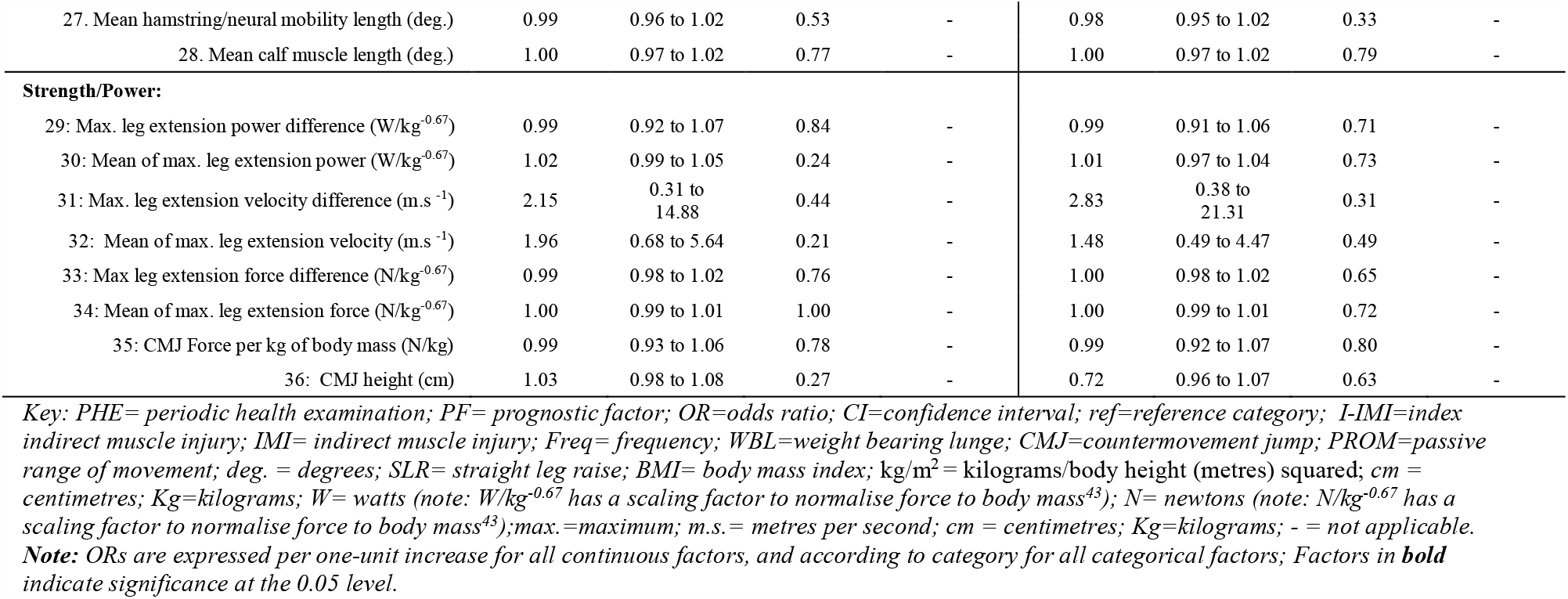
Univariable and multivariable logistic regression estimates for all candidate prognostic factors

| Candidate PF & Type | Univariable (unadjusted) |  |  |  | Multivariable (adjusted for age, height, weight) |  |  |  |
| --- | --- | --- | --- | --- | --- | --- | --- | --- |
|  | OR | 95% CI | P | Best model fit | OR | 95% CI | P | Best model fit |
| **Anthropometric (Adjustment Factors):** |  |  |  |  |  |  |  |  |
| 1: Age (years) | **1.12** | **1.06-1.18** | **<0.001** | **Linear** | **1.12** | **1.05 to 1.18** | **<0.001** | **Linear** |
| 2: Height (cm) | 1.03 | 0.99 to 1.07 | 0.13 | - | 1.02 | 0.97 to 1.07 | 0.46 | - |
| 3: Weight (kg) | **1.03** | **1.00 to 1.07** | **0.03** | **Linear** | 1.00 | 0.95 to 1.04 | 0.91 | - |
| **Within 3 years preceding PHE, frequency of:** |  |  |  |  |  |  |  |  |
| 4: foot/ankle injuries | 1.04 | 0.87 to 1.24 | 0.68 | - | 1.04 | **0.87 to 1.25** | 0.65 | - |
| 5: hip/groin injuries | 1.16 | 0.90 to 1.51 | 0.25 | - | 1.29 | 0.99 to 1.70 | 0.06 | - |
| 6: knee injuries | 0.96 | 0.72 to 1.29 | 0.81 | - | 0.98 | 0.72 to 1.32 | 0.88 | - |
| 7: shoulder injuries | 2.38 | 0.98 to 5.75 | 0.05 | - | 1.89 | 0.76 to 4.74 | 0.17 | - |
| 8: lumbar spine injuries | 0.88 | 0.60 to 1.29 | 0.50 | - | 1.08 | 0.72 to 1.61 | 0.71 | - |
| 9: iliopsoas IMIs | 0.73 | 0.38 to 1.43 | 0.37 | - | 0.86 | 0.43 to 1.71 | 0.67 | - |
| 10: hip adductor IMIs | 1.38 | 0.92 to 2.09 | 0.12 | - | 1.18 | 0.76 to 1.84 | 0.46 | - |
| 11: hamstring IMIs | **1.56** | **1.17 to 2.09** | **<0.001** | **Linear** | 1.35 | 1.00 to 1.83 | 0.05 | - |
| 12: quadriceps IMIs | 1.08 | 0.67 to 1.73 | 0.75 | - | 1.05 | 0.65 to 1.71 | 0.84 | - |
| 13: calf IMIs | **1.80** | **1.09 to 2.97** | **0.02** | **Linear** | 1.30 | 0.75 to 2.25 | 0.35 | - |
| **Within 3 years preceding PHE, most recent:** |  |  |  |  |  |  |  |  |
| 14: foot/ankle injury (never) | ref | ref | ref | - | ref | ref | ref | - |
| 14: foot/ankle injury (<6 months) | 1.27 | 0.64 to 2.53 | 0.49 | - | 1.41 | 0.69 to 2.87 | 0.35 | - |
| 14: foot/ankle injury (6-12 months) | 1.16 | 0.54 to 2.46 | 0.71 | - | 1.15 | 0.52 to 2.52 | 0.73 | - |
| 14: foot/ankle injury (>12 months) | 1.27 | 0.75 to 2.13 | 0.37 | - | 1.22 | 0.71 to 2.09 | 0.47 | - |
| 15: hip/groin injury (never) | ref | ref | ref | - | ref | ref | ref | - |
| 15: hip/groin injury (<6 months) | 1.05 | 0.44 to 2.49 | 0.92 | - | 1.66 | 0.67 to 4.13 | 0.27 | - |
| 15: hip/groin injury (6-12 months) | 0.59 | 0.23 to 1.50 | 0.27 | - | 0.65 | 0.25 to 1.69 | 0.38 | - |
| 15: hip/groin injury (>12months) | 1.58 | 0.87 to 2.87 | 0.14 | - | 1.80 | 0.96 to 3.36 | 0.07 | - |
| 16: knee injury (never) | ref | ref | ref | - | ref | ref | ref | - |
| 16: knee injury (<6 months) | 1.15 | 0.40 to 3.28 | 0.80 | - | 1.12 | 0.37 to 3.34 | 0.84 | - |
| 16: knee injury (6-12 months) | 1.23 | 0.58 to 2.62 | 0.60 | - | 1.24 | 0.57 to 2.70 | 0.60 | - |
| 16: knee injury (>12months) | 0.93 | 0.53 to 1.61 | 0.79 | - | 0.99 | 0.56 to 1.75 | 0.96 | - |
| 17: shoulder injury (never) | ref | ref | ref | - | ref | ref | ref | - |
| 17: shoulder injury (<6 months) | 2.75 | 0.50 to 15.26 | 0.25 | - | 2.55 | 0.44 to 14.72 | 0.30 | - |
| 17: shoulder injury (6-12 months) | 1.38 | 0.19 to 9.90 | 0.75 | - | 1.11 | 1.15 to 8.36 | 0.92 | - |
| 17: shoulder injury (>12months) | 3.21 | 0.81 to 12.67 | 0.10 | - | 2.36 | 0.58 to 9.62 | 0.23 | - |
| 18: lumbar spine injury (never) | ref | ref | ref | - | ref | ref | ref | - |
| 18: lumbar spine injury (<6 months) | 1.24 | 0.30 to 5.05 | 0.77 | - | 2.10 | 0.50 to 8.82 | 0.31 | - |
| 18: lumbar spine injury (6-12 months) | 0.62 | 0.15 to 2.53 | 0.50 | - | 0.75 | 0.18 to 3.14 | 0.69 | - |
| 18: lumbar spine injury (>12months) | 0.70 | 0.34 to 1.44 | 0.33 | - | 0.97 | 0.46 to 2.06 | 0.94 | - |
| 19: iliopsoas IMI (never) | ref | ref | ref | - | ref | ref | ref | - |
| 19: iliopsoas IMI (<6 months) | 1.24 | 0.77 to 20.05 | 0.88 | - | 1.27 | 0.08 to 21.32 | 0.87 | - |
| 19: iliopsoas IMI (6-12 months) | 0.62 | 0.15 to 2.53 | 0.51 | - | 0.83 | 0.20 to 3.43 | 0.80 | - |
| 19: iliopsoas IMI (>12months) | 0.57 | 0.21 to 1.55 | 0.27 | - | 0.66 | 0.24 to 1.82 | 0.42 | - |
| 20: hip adductor IMI (never) | ref | ref | ref | - | ref | ref | ref | - |
| 20: hip adductor IMI (<6 months) | 1.37 | 0.53 to 3.56 | 0.52 | - | 1.29 | 0.48 to 3.47 | 0.61 | - |
| 20: hip adductor IMI (6-12 months) | 1.37 | 0.43 to 4.36 | 0.60 | - | 1.16 | 0.35 to 3.81 | 0.81 | - |
| 20: hip adductor IMI (>12months) | 1.37 | 0.59 to 3.16 | 0.46 | - | 1.14 | 0.47 to 2.79 | 0.77 | - |
| 21: hamstring IMI (never) | ref | ref | ref | - | ref | ref | ref | - |
| 21: hamstring IMI (<6 months) | 2.84 | 0.81 to 9.99 | 0.10 | - | 2.07 | 0.57 to 7.56 | 0.27 | - |
| 21: hamstring IMI (6-12 months) | 1.42 | 0.66 to 3.06 | 0.37 | - | 1.22 | 0.56 to 2.70 | 0.62 | - |
| 21: hamstring IMI (>12months) | **2.95** | **1.51 to 5.73** | **<0.001** | **Linear** | **2.24** | **1.11 to 4.53** | **0.02** | **Linear** |
| 22: quadriceps IMI (never) | ref | ref | ref | - | ref | ref | ref | - |
| 22: quadriceps IMI (<6 months) | 1.74 | 0.38 to 7.91 | 0.48 | - | 1.56 | 0.33 to 7.36 | 0.58 | - |
| 22: quadriceps IMI (6-12 months) | 0.58 | 0.17 to 1.93 | 0.37 | - | 0.59 | 0.17 to 2.04 | 0.41 | - |
| 22: quadriceps IMI (>12months) | 1.14 | 0.53 to 2.43 | 0.74 | - | 1.07 | 0.49 to 2.35 | 0.86 | - |
| 23: calf IMI (never) | ref | ref | ref | - | ref | ref | ref | - |
| 23: calf IMI (<6 months) | 3.78 | 0.98 to 14.56 | 0.05 | - | 3.11 | 0.78 to 12.46 | 0.11 | - |
| 23: calf IMI (6-12 months) | 7.09 | 0.82 to 61.51 | 0.08 | - | 3.80 | 0.41 to 35.41 | 0.24 | - |
| 23: calf IMI (>12months) | 1.26 | 0.47 to 3.36 | 0.64 | - | 0.73 | 0.24 to 2.15 | 0.56 | - |
| **Musculoskeletal:** |  |  |  |  |  |  |  |  |
| 24. Mean PROM hip IR (deg.) | **0.97** | **0.95 to 0.99** | **0.01** | **Linear** | 0.98 | 0.95 to 1.00 | 0.06 | - |
| 25. Mean PROM hip ER (deg.) | 0.97 | 0.95 to 1.00 | 0.09 | - | 0.99 | 0.96 to 1.02 | 0.53 | - |
| 26. Mean hip flexor length (deg.) | 1.01 | 0.98 to 1.04 | 0.46 | - | 1.01 | 0.99 to 1.04 | 0.32 | - |
| 27. Mean hamstring/neural mobility length (deg.) | 0.99 | 0.96 to 1.02 | 0.53 | - | 0.98 | 0.95 to 1.02 | 0.33 | - |
| 28. Mean calf muscle length (deg.) | 1.00 | 0.97 to 1.02 | 0.77 | - | 1.00 | 0.97 to 1.02 | 0.79 | - |
| **Strength/Power:** |  |  |  |  |  |  |  |  |
| 29: Max. leg extension power difference (W/kg<sup>-0.67</sup>) | 0.99 | 0.92 to 1.07 | 0.84 | - | 0.99 | 0.91 to 1.06 | 0.71 | - |
| 30: Mean of max. leg extension power (W/kg<sup>-0.67</sup>) | 1.02 | 0.99 to 1.05 | 0.24 | - | 1.01 | 0.97 to 1.04 | 0.73 | - |
| 31: Max. leg extension velocity difference (m.s<sup>-1</sup>) | 2.15 | 0.31 to 14.88 | 0.44 | - | 2.83 | 0.38 to 21.31 | 0.31 | - |
| 32: Mean of max. leg extension velocity (m.s<sup>-1</sup>) | 1.96 | 0.68 to 5.64 | 0.21 | - | 1.48 | 0.49 to 4.47 | 0.49 | - |
| 33: Max leg extension force difference (N/kg<sup>-0.67</sup>) | 0.99 | 0.98 to 1.02 | 0.76 | - | 1.00 | 0.98 to 1.02 | 0.65 | - |
| 34: Mean of max. leg extension force (N/kg<sup>-0.67</sup>) | 1.00 | 0.99 to 1.01 | 1.00 | - | 1.00 | 0.99 to 1.01 | 0.72 | - |
| 35: CMJ Force per kg of body mass (N/kg) | 0.99 | 0.93 to 1.06 | 0.78 | - | 0.99 | 0.92 to 1.07 | 0.80 | - |
| 36: CMJ height (cm) | 1.03 | 0.98 to 1.08 | 0.27 | - | 0.72 | 0.96 to 1.07 | 0.63 | - |

Key: PHE= periodic health examination; PF= prognostic factor; OR=odds ratio; CI=confidence interval; ref=reference category; I-IMI=index indirect muscle injury; IMI= indirect muscle injury; Freq= frequency; WBL=weight bearing lunge; CMJ=countermovement jump; PROM=passive range of movement; deg. = degrees; SLR= straight leg raise; BMI= body mass index; kg/m<sup>2</sup>= kilograms/body height (metres) squared; cm = centimetres; Kg=kilograms; W= watts (note: W/kg<sup>-0.67</sup> has a scaling factor to normalise force to body mass<sup>43</sup>); N= newtons (note: N/kg<sup>-0.67</sup> has a scaling factor to normalise force to body mass<sup>43</sup>); max.=maximum; m.s.= metres per second; cm = centimetres; Kg=kilograms; - = not applicable. **Note:** ORs are expressed per one-unit increase for all continuous factors, and according to category for all categorical factors; Factors in **bold** indicate significance at the 0.05 level.

Despite relatively large ORs, weaker evidence of associations was observed for the frequency of previous shoulder injuries (OR=2.38, 95% CI = 0.98 to 5.75, p=0.05) and if the most recent calf IMI was less than 6 months prior to PHE (OR=3.78, 95% CI=0.98 to 14.56, p=0.05). However, the very wide CIs indicated considerable uncertainty about the true OR. No other significant candidate factors were observed.

### Multivariable analyses

Table 4 shows the results of the multivariable analyses, where the adjusted prognostic value was evaluated for all PHE-derived variables. After adjustment for height and weight, age remained significantly associated with increased odds of sustaining an I-IMI during a season (OR=1.12 for a 1-year increase, 95%CI = 1.05 to 1.18, p<0.001) and a linear association was the best fit for this variable. However, there was no evidence that height and weight were strong prognostic factors independent of age.

After adjustment for age, height and weight, if the most recent hamstring IMI was more than 12 months but less than 3 years prior to PHE, the significant association and wide CI also remained (OR= 2.24, 95% CI = 1.11 to 4.53, p=0.02). However, no other candidates demonstrated prognostic importance. For most variables, the magnitude of the adjusted prognostic association was also smaller than the unadjusted association and some confidence intervals were very wide.

### Complete case and sensitivity analysis

The results of all complete case and sensitivity analyses are presented in supplementary files 5-7). Figures 2 and 3 show forest plots of the estimates obtained for all statistically significant candidate prognostic factors across all univariable and multivariable analyses.

**Figure 2:**
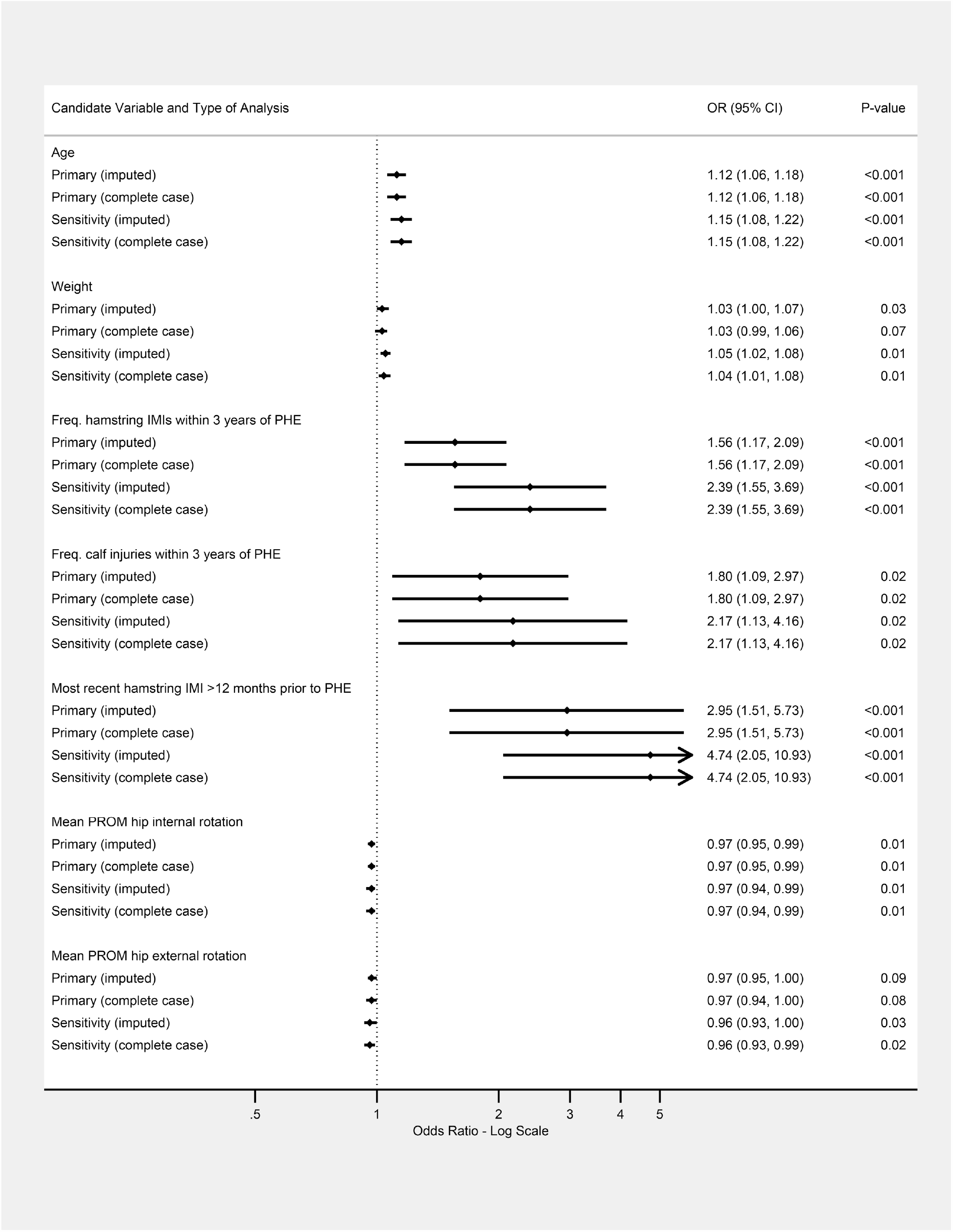
Comparison of candidate prognostic factors with significant associations across the primary and sensitivity univariable analyses (using imputed and complete case datasets). *Key: OR = odds ratio; freq. = frequency; IMI = indirect muscle injury; PHE = periodic health examination; PROM = passive range of movement*.

**Figure 3:**
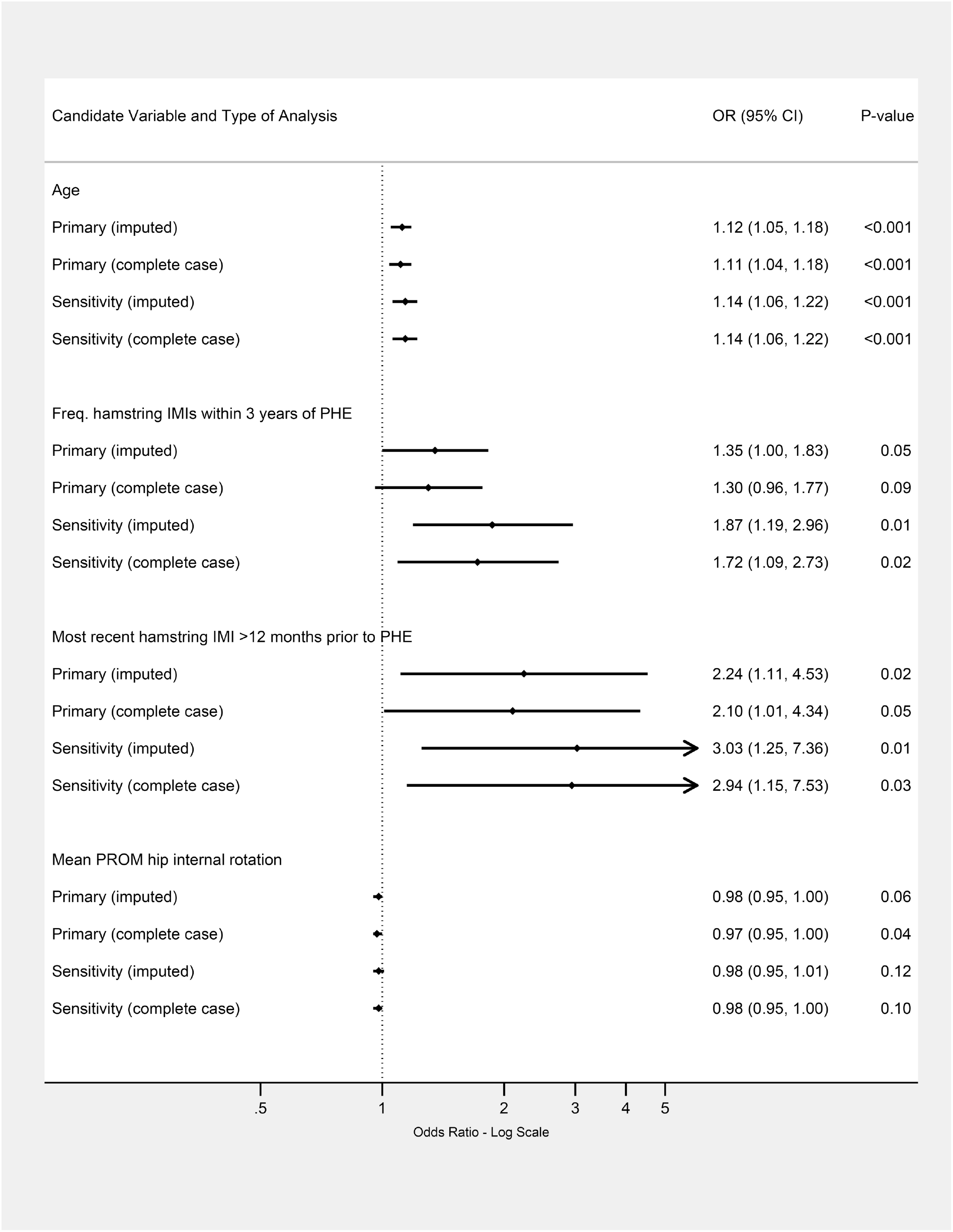
Comparison of candidate prognostic factors with significant associations across the primary and sensitivity multivariable analyses (using imputed and complete case datasets) *Key: OR = odds ratio; freq. = frequency; IMI = indirect muscle injury; PHE = periodic health examination; PROM = passive range of movement. Note: the factor of age was adjusted for height and weight. All other factors were adjusted for age, height and weight*.

For both univariable and multivariable analyses, the prognostic associations were very similar for the complete case and imputation analyses. Sensitivity analyses (that is, excluding players who were loaned or transferred) led to slightly stronger prognostic associations for some factors (for example, the frequency and timing of previous hamstring injuries), although the CIs were wider.

## DISCUSSION

This exploratory study has examined linear and non-linear prognostic associations between PHE-derived variables and I-IMIs in elite football players, using routinely collected data from a 5-season period.

While the univariable analyses suggested 6 candidate factors were associated with I-IMIs, such analyses are limited as they only provide baseline association measures prior to adjustment for other important prognostic factors.^29^ However, after adjustment for age, height and weight in the multivariable analyses, we found that most PHE derived variables did not add any prognostic value over and above age. The exception was that if a hamstring IMI was sustained by a player more than 12 months (but less than 3 years) prior to PHE, their odds of sustaining a lower extremity I-IMI significantly increased 2.2-fold, which has not been previously reported. Although not directly comparable, earlier studies have also shown that a history of a previous hamstring IMI is specifically associated with an increased hazard of future hamstring IMIs in elite players.^11 20 22^ Nevertheless, the uncertainty in our estimates (demonstrated by wide 95% confidence intervals) and differences observed during the sensitivity analyses mean that this variable only has provisional prognostic value and needs to be established in further confirmatory studies.

Indeed, age was the only variable that could be considered as an important prognostic factor, which is easily obtained even without conducting PHE; after allowing for height and weight, age retained a highly significant, modest and precise association that was robust to both imputation and player transfers. For illustration, our estimates suggest that for every 1-year increase in age, the odds of sustaining an I-IMI during a season would increase by approximately 12%. As an example (to put into the context of absolute risk), for two players who were the same height and weight but aged 5 years apart, if the younger-aged player had a risk of 0.44 (which was overall outcome prevalence in our study), then the older player would have a risk of 0.58.

Our findings confirm those of our multivariable prognostic model study, where age was retained in a model to determine individualised I-IMI risk over a season in elite players (OR = 1.10, 95% CI = 1.03 to 1.17).^17^ Other studies have shown that as a multivariable prognostic factor, age is associated specifically with increased hamstring IMI risk (OR range 1.40-1.78), 11 18 23 where the reported estimates were larger than those observed in our study. These differences may be due to chance or partly because we merged all lower extremity I-IMI outcomes rather than utilising IMI subgroups, which may have diluted the strength of our observed associations. However, although our approach was less clinically meaningful, merging I-IMI outcomes was essential in order to maximise the statistical power of our study.

Importantly, while multiple exploratory studies have confirmed the prognostic importance of age,^11 17 18 23^ it is not a causal factor for future IMI occurrence. Rather, it is likely to be a proxy marker for another potential causal mechanism, although to establish the true causal role of a prognostic factor, associations should be demonstrated during additional confirmatory and explanatory studies.^50^ Even taking this into account, injury prevention strategies have been traditionally developed to modify supposed causal factors,^6^ but because age is non-modifiable it could not be used to inform this process in a clinical setting. Instead, age should currently only be considered clinically useful to explain differences in risk between players in a team, or included as an important factor in future prognostic model development studies.

Using data from PHE tests that measure modifiable physical and performance characteristics has been previously questioned for injury prediction purposes.^2^ Our results fully support this view, because none of the modifiable musculoskeletal (clinical examination) or strength and power tests evaluated showed any statistically significant associations with I-IMIs. This absence of strong associations mean that such tests have poor discriminatory ability, usually because of overlap in test scores that occur in individuals who sustain a future injury and those who do not.^2^ Furthermore, after measurement at a solitary timepoint (i.e. pre-season), it is likely that the prognostic value of modifiable factors is time-varying^51^ as a consequence of physical and physiological adaptations that occur from training exposure and other injuries.^52^

Overall, our findings suggest that at present, PHE cannot be considered a useful source of prognostic factors for IMI risk prediction and injury prevention practice in elite football players. Instead, PHE should only be deemed clinically useful for screening of pathology that could pose a risk to safe participation and for rehabilitation or performance monitoring purposes. However, because of our study’s exploratory nature (with many estimates having very wide confidence intervals), the shortcomings of the current evidence base and the paucity of known prognostic factors in elite football,^11^ there is a clear need for further investigation in this area to improve our understanding of the prognostic value of PHE in elite football and other sports.

### Limitations and future research

Our study is unique in that we have investigated non-linear associations as per methodological guidelines.^9 53^ However, in our analyses linear associations were selected over non-linear associations. For practical reasons, our imputation did not assume non-linear associations and therefore may have reduced our ability to detect genuine non-linear relationships in the subsequent analyses. However, this is not a concern for age, as there were no missing values for this factor and is unlikely to be a material concern for all other factors as missing data was always less than 15%.

A competing risks analysis was not conducted, which meant that individuals who sustained injury types other than lower extremity IMIs were still considered at risk, even though this may have affected their training and match exposure and hence risk of sustaining an I-IMI event. We also only measured candidate factors at one timepoint each season, which means that dynamic associations were not investigated. We also assumed that participant-seasons were independent. Future studies could account for competing risks, utilise repeated measurements over time and incorporate between-season correlations into analyses. However, the complexity such analyses would also require a significantly larger volume of data. This could be achieved through data sharing initiatives and individual participant data meta-analysis, which would also increase the power to detect genuine prognostic associations and non-linear relationships.

## CONCLUSION

We have evaluated prognostic associations between PHE-derived candidate variables and lower extremity I-IMIs in elite football players using data that were routinely collected over 5 seasons. We found no clear associations for nearly all PHE variables, although if a player sustained a hamstring IMI greater than 12 months (but less than 3 years) prior to PHE, then this had potential prognostic value over and above the prognostic value of age. Indeed, age was the only variable to be confirmed as a clear prognostic factor in both univariable and multivariable analyses. However, this is easily measured without then need to conduct PHE, and although it has limited application in clinical practice, it should be included as an important factor in any future prognostic model development studies. Our findings suggest that overall, PHE cannot be currently considered as a useful source of prognostic factors for I-IMI risk prediction and injury prevention practice in elite football players. Instead, it should only be utilised for screening of pathology, as well as rehabilitation and performance monitoring. Further research is required to confirm the prognostic value of PHE for IMI risk prediction and to identify novel prognostic factors that could improve development of prognostic models in football and other sports.

## Supporting information

REMARK CHECKLIST

## Data Availability

An anonymised summary of the dataset that was analysed during this study may be available from the corresponding author on reasonable request.

## ACKNOWLEDGEMENTS

The authors would like to thank all staff within the Medical and Sports Science Department at Manchester United for their continuing help and support with this manuscript and thank all players for their participation (without whom this study would not be possible). The authors also thank Versus Arthritis for their support: Versus Arthritis grant number 21755.

## CONTRIBUTORS

TH was responsible for the conceptualisation of the project, study design, database construction, data extraction and cleaning, protocol development and protocol writing. TH conducted the data analysis, interpretation and wrote the main manuscript. RR provided statistical guidance and assisted with development of the study design, analysis, and edited manuscript drafts. MC assisted with the study conceptualisation and design, protocol development, clinical interpretation and editing the manuscript drafts. JS provided guidance with the study design, development of the analysis and protocol, interpretation of the analysis, as well as editing the study manuscripts. All authors read and approved this final manuscript.

## ETHICS AND DATA USE

Informed consent was not required as data were captured from the mandatory PHE completed through the participants’ employment. The data usage was approved by the football club and the Research Ethics Service at the University of Manchester.

## PATIENT CONSENT FOR PUBLICATION

Not required.

