## SUPPLEMENTARY FILE 1 for "Can prognostic factors for indirect muscle injuries in elite football (soccer) players be identified using data from preseason screening? An exploratory analysis using routinely-collected periodic health examination records"

**Supplementary file 1: Sample size calculation**

PASS 2019, v19.0.2	13/11/2019 14:40:18	1

**Tests for the Odds Ratio in Logistic Regression with One Normal X and Other Xs (Wald Test)**

**Numeric Results ────────────────────────────────────────────────────────────**

**Odds	R**

**Power	N	P0	P1	Ratio	Squared	Alpha	Beta**

0.79904	314	0.430	0.542	1.570	0.500	0.050	0.20096

Logistic Regression', Statistics in Medicine, Volume 17, pages 1623-1634.

**Report Definitions**

Power is the probability of rejecting a false null hypothesis. It should be close to one.

N is the size of the sample drawn from the population.

P0 is the response probability at the mean of X.

P1 is the response probability when X is increased to one standard deviation above the mean.

Odds Ratio is the odds ratio when P1 is on top. That is, it is [P1/(1-P1)]/[P0/(1-P0)].

R-Squared is the R2 achieved when X is regressed on the other independent variables in the regression.

Alpha is the probability of rejecting a true null hypothesis.

Beta is the probability of accepting a false null hypothesis.

**Summary Statements ─────────────────────────────────────────────────────────**

A logistic regression of a binary response variable (Y) on a continuous, normally distributed

variable (X) with a sample size of 314 observations achieves 80% power at a 0.050 significance

level to detect a change in Prob(Y=1) from the value of 0.430 at the mean of X to 0.542 when X

is increased to one standard deviation above the mean. This change corresponds to an odds ratio

of 1.570. An adjustment was made since a multiple regression of the independent variable of

interest on the other independent variables in the logistic regression obtained an R-Squared of

0.500.

**Dropout-Inflated Sample Size ────────────────────────────────────────────────────**

**Dropout-**

**Inflated	Expected**

**Enrollment	Number of**

**Sample Size	Sample Size	Dropouts**

**Dropout Rate	N	N'	D**

20%	314	393	79

**Definitions**

Dropout Rate (DR) is the percentage of subjects (or items) that are expected to be lost at random during the

course of the study and for whom no response data will be collected (i.e. will be treated as "missing").

N is the evaluable sample size at which power is computed. If N subjects are evaluated out of the N' subjects

that are enrolled in the study, the design will achieve the stated power.

N' is the total number of subjects that should be enrolled in the study in order to end up with N evaluable

subjects, based on the assumed dropout rate. After solving for N, N' is calculated by inflating N using the

formula N' = N / (1 - DR), with N' always rounded up. (See Julious, S.A. (2010) pages 52-53, or Chow, S.C.,

Shao, J., Wang, H., and Lokhnygina, Y. (2018) pages 32-33.)

D is the expected number of dropouts. D = N' - N.

PASS 2019, v19.0.2	13/11/2019 14:40:18	2

**Tests for the Odds Ratio in Logistic Regression with One Normal X and Other Xs (Wald Test)**

**Procedure Input Settings ──────────────────────────────────────────────────────**

Autosave Inactive

**Design Tab**

Solve For:	Sample Size

Alternative Hypothesis:	Two-Sided

Power:	0.80

Alpha:	0.05

P0 (Baseline Probability that Y=1):	0.43

Use P1 or Odds Ratio:	Odds Ratio

Odds Ratio (Odds1/Odds0):	1.57

R-Squared of X1 with Other X's:	0.5
