## SUPPLEMENTARY FILE 2 for "Can prognostic factors for indirect muscle injuries in elite football (soccer) players be identified using data from preseason screening? An exploratory analysis using routinely-collected periodic health examination records"

**Supplementary file 2: Candidate prognostic factors that were excluded**

| Type of prognostic factor | Candidate Prognostic Factor | Composite variable created | Measurement unit | Measurement method | Data type | Reason for elimination |
| --- | --- | --- | --- | --- | --- | --- |
| Anthropometric | Body fat | No | Percentage | Skin calipers | Continuous | Missing data > 15% |
| Musculoskeletal test | Quadriceps muscle length difference* | Yes | Degrees | Ely’s test using digital inclinometer | Continuous | Intra-rater ICC = 0.69^1^  Inter-rater ICC = 0.66. ^1^ |
|  | Mean quadriceps muscle length** | Yes | Degrees | Ely’s test using goniometer | Continuous | Intra-rater ICC = 0.69^1^  Inter-rater ICC = 0.66. ^1^ |
|  | Toe touch in standing | No | Centimetres | Fingertips to floor distance | Continuous | Missing data > 15% |
|  | Sacroiliac joint kinematic function | No | Subjective score | Gillets test | Categorical | Missing data > 15% |
| Functional movement/balance tests | Y Balance Test – anterior translation difference* | Yes | Centimetres | Y Balance Test | Continuous | Missing data >15% |
|  | Y Balance Test – Mean anterior translation** | Yes | Centimetres | Y Balance Test | Continuous | Missing data >15% |
|  | Y Balance Test – posteromedial translation difference* | Yes | Centimetres | Y Balance Test | Continuous | Missing data >15% |
|  | Y Balance Test – Mean posteromedial translation** | Yes | Centimetres | Y Balance Test | Continuous | Missing data >15% |
|  | Y Balance Test –posterolateral translation difference* | Yes | Centimetres | Y Balance Test | Continuous | Missing data >15% |
|  | Y Balance Test – Mean posterolateral translation** | Yes | Centimetres | Y Balance Test | Continuous | Missing data >15% |
|  | R relative tibial angles | No | Degrees | SLS measurement with Dorsavi Viperform IMU | Continuous | Within-session ICCs=0.27-0.75  Between-session ICCs = 0.55-0.77^2^ |
|  | L relative tibial angles (left leg) | No | Degrees | SLS measurement with Dorsavi Viperform IMU | Continuous | Within-session ICCs= 0.27-0.75  Between-session ICCs = 0.55-0.77^2^ |
| Strength/power tests | Upper body peak power difference* | Yes | Normalised watts per kilo (W/kg^-0.67^) | Double horizontal press using a Keiser Chest Press Air 350 machine | Continuous | Missing data > 15% |
|  | Mean upper body peak power** | Yes | Normalised watts per kilo (W/kg^-0.67^) | Double horizontal press using a Keiser Chest Press Air 350 machine | Continuous | Missing data > 15% |

*Key: PHE=periodic health examination; WBL=weight bearing lunge; CMJ=countermovement jump; PROM=passive range of movement; ICC=intraclass correlation coefficient; SLR= straight leg raise; SLS = single leg squat; BMI= body mass index; f=force; W= watts; (note that W/kg^-0.67^ has a scaling factor to normalise power to body mass) Kg=kilos; IMU= inertial measurement units; m = mass; Note: composite factors are identified in the table with * denoting between limb differences and **denoting combined mean values of both limbs.*
