## SUPPLEMENTARY FILE 3 for "Can prognostic factors for indirect muscle injuries in elite football (soccer) players be identified using data from preseason screening? An exploratory analysis using routinely-collected periodic health examination records"

**Supplementary file 3: Anthropometric parameters and all included candidate prognostic factor characteristics for participants included in the sensitivity analyses**

| ***Characteristic/candidate prognostic factors*** | | ***Min*** | ***Lower quartile*** | ***Median*** | ***Upper quartile*** | ***Max.*** | ***Freq. (%) - if categorical*** | ***Missing values n (%)*** |
| --- | --- | --- | --- | --- | --- | --- | --- | --- |
| **Anthropometrics** |  |  |  |  |  |  |  |  |
| 1. Age at PHE (years) |  | 16.01 | 17.64 | 19.20 | 24.30 | 39.59 | - | 0 (0) |
| 2. Height (cm) |  | 164.30 | 176.10 | 180.00 | 185.50 | 195.00 | - | 15 (5.77) |
| 3. Weight (kg) |  | 56.80 | 68.09 | 73.10 | 80.00 | 94.00 | - | 16 (6.15) |
| **Past medical history** |  |  |  |  |  |  |  |  |
| Within 3 years prior to PHE, freq. of: |  |  |  |  |  |  |  |  |
| 4. foot/ankle injuries |  | 0.00 | 0.00 | 1.00 | 2.00 | 7.00 | - | 0 (0) |
| 5. Hip/groin injuries |  | 0.00 | 0.00 | 0.00 | 1.00 | 5.00 | - |  |
| 6. Knee injuries |  | 0.00 | 0.00 | 0.00 | 1.00 | 3.00 | - | 0 (0) |
| 7. Shoulder injuries |  | 0.00 | 0.00 | 0.00 | 0.00 | 2.00 | - | 0 (0) |
| 8. lumbar spine injuries |  | 0.00 | 0.00 | 0.00 | 0.00 | 3.00 | - | 0 (0) |
| 9. iliopsoas IMIs |  | 0.00 | 0.00 | 0.00 | 0.00 | 2.00 | - | 0 (0) |
| 10. hip adductor IMIs |  | 0.00 | 0.00 | 0.00 | 0.00 | 3.00 | - | 0 (0) |
| 11.hamstring IMIs |  | 0.00 | 0.00 | 0.00 | 1.00 | 5.00 | - | 0 (0) |
| 12. quadriceps IMIs |  | 0.00 | 0.00 | 0.00 | 0.00 | 3.00 | - | 0 (0) |
| 13. calf IMIs |  | 0.00 | 0.00 | 0.00 | 0.00 | 4.00 | - | 0 (0) |
| **Within 3 years prior to PHE, most recent:** |  |  |  |  |  |  |  |  |
| 14. foot/ankle injury | *Never* | - | - | - | - | - | 120 (46.15) | 0 (0) |
|  | *< 6 months* | - | - | - | - | - | 37 (14.23) | 0 (0) |
|  | *6-12 months* | - | - | - | - | - | 29 (11.15) | 0 (0) |
|  | *>12 months* | - | - | - | - | - | 74 (28.46) | 0 (0) |
| 15. Hip/groin injury | *Never* | - | - | - | - | - | 176 (67.69) | 0 (0) |
|  | *< 6 months* | - | - | - | - | - | 21 (8.08) | 0 (0) |
|  | *6-12 months* | - | - | - | - | - | 20 (7.69) | 0 (0) |
|  | *>12 months* | - | - | - | - | - | 43 (16.54) | 0 (0) |
| 16. Knee injury | *Never* | - | - | - | - | - | 165 (63.46) | 0 (0) |
|  | *< 6 months* | - | - | - | - | - | 15 (5.77) | 0 (0) |
|  | *6-12 months* | - | - | - | - | - | 24 (9.23) | 0 (0) |
|  | *>12 months* | - | - | - | - | - | 56 (21.54) | 0 (0) |
| 17. Shoulder injury | *Never* | - | - | - | - | - | 240 (92.31) | 0 (0) |
|  | *< 6 months* | - | - | - | - | - | 6 (2.31) | 0 (0) |
|  | *6-12 months* | - | - | - | - | - | 4 (1.54) | 0 (0) |
|  | *>12 months* | - | - | - | - | - | 10 (3.85) | 0 (0) |
| 18. lumbar spine injury | *Never* | - | - | - | - | - | 210 (80.77) | 0 (0) |
|  | *< 6 months* | - | - | - | - | - | 8 (3.08) | 0 (0) |
|  | *6-12 months* | - | - | - | - | - | 7 (2.69) | 0 (0) |
|  | *>12 months* | - | - | - | - | - | 35 (13.46) | 0 (0) |
| 19. iliopsoas IMI | *Never* | - | - | - | - | - | 237 (91.15) | 0 (0) |
|  | *< 6 months* | - | - | - | - | - | 2 (0.77) | 0 (0) |
|  | *6-12 months* | - | - | - | - | - | 7 (2.69) | 0 (0) |
|  | *>12 months* | - | - | - | - | - | 14 (5.38) | 0 (0) |
| 20. hip adductor IMI | *Never* | - | - | - | - | - | 214 (82.31) | 0 (0) |
|  | *< 6 months* | - | - | - | - | - | 17 (6.54) | 0 (0) |
|  | *6-12 months* | - | - | - | - | - | 10 (3.85) | 0 (0) |
|  | *>12 months* | - | - | - | - | - | 19 (7.31) | 0 (0) |
| 21.hamstring muscle IMI | *Never* | - | - | - | - | - | 193 (74.23) | 0 (0) |
|  | *< 6 months* | - | - | - | - | - | 8 (3.08) | 0 (0) |
|  | *6-12 months* | - | - | - | - | - | 23 (8.85) | 0 (0) |
|  | *>12 months* | - | - | - | - | - | 36 (13.85) | 0 (0) |
| 22. quadriceps IMI | *Never* | - | - | - | - | - | 165 (63.46) | 0 (0) |
|  | *< 6 months* | - | - | - | - | - | 5 (1.92) | 0 (0) |
|  | *6-12 months* | - | - | - | - | - | 12 (4.62) | 0 (0) |
|  | *>12 months* | - | - | - | - | - | 24 (9.23) | 0 (0) |
| 23. calf IMI | *Never* | - | - | - | - | - | 230 (88.46) | 0 (0) |
|  | *< 6 months* | - | - | - | - | - | 10 (3.85) | 0 (0) |
|  | *6-12 months* | - | - | - | - | - | 5 (1.92) | 0 (0) |
|  | *>12 months* | - | - | - | - | - | 15 (5.77) | 0 (0) |
| **Musculoskeletal Examination** |  |  |  |  |  |  |  |  |
| 24. Mean PROM hip IR (deg.) |  | 9.50 | 22.00 | 32.50 | 40.00 | 55.00 | - | 15 (5.77) |
| 25. Mean PROM hip ER (deg.) |  | 17.50 | 33.70 | 39.00 | 43.50 | 62.00 | - | 15 (5.77) |
| 26. Mean hip flexor length (deg.) |  | -7.00 | 3.00 | 8.68 | 15.00 | 55.00 | - | 18 (6.92) |
| 27. Mean hamstring/neural mobility length (deg.) |  | 45.00 | 83.00 | 90.00 | 90.00 | 102.00 | - | 15 (5.77) |
| 28. Mean calf muscle length (deg.) |  | 9.50 | 25.65 | 30.50 | 37.00 | 57.50 | - | 15 (5.77) |
| **Strength/Power Tests** |  |  |  |  |  |  |  |  |
| 29: Max. leg extension power difference (W/kg^-0.67^) |  | -11.94 | -1.47 | 0.43 | 1.91 | 15.26 | - | 39 (15.00) |
| 30: Mean of max. leg extension power (W/kg^-0.67^) |  | 23.01 | 41.67 | 46.40 | 51.10 | 78.70 | - | 39 (15.00) |
| 31: Max. leg extension velocity difference (m.s ^-1^) |  | -0.34 | -0.05 | 0.02 | 0.11 | 0.40 | - | 35 (13.46) |
| 32: Mean of max. leg extension velocity (m.s ^-1^) |  | 1.02 | 1.68 | 1.84 | 1.98 | 2.35 | - | 35 (13.46) |
| 33: Max leg extension force difference (N/kg^-0.67^) |  | -83.59 | -5.74 | -1.07 | 4.39 | 55.41 | - | 39 (15.00) |
| 34: Mean of max. leg extension force (N/kg^-0.67^) |  | 50.19 | 90.85 | 101.14 | 112.02 | 217.96 | - | 39 (15.00) |
| 35: CMJ Force per kg of body mass |  | 20.50 | 23.30 | 25.18 | 27.90 | 39.20 | - | 35 (13.46) |
| 36: CMJ height (cm) |  | 28.70 | 37.20 | 39.90 | 43.10 | 58.00 | - | 35 (13.46) |

*Key: PHE= periodic health examination; I-IMI=index indirect muscle injury; IMI= indirect muscle injury; min = minimum; max = maximum; n = observations; Freq= frequency; WBL=weight bearing lunge; CMJ=countermovement jump; PROM=passive range of movement; deg. = degrees; SLR= straight leg raise; BMI= body mass index;* kg/m^2^ = kilograms/body height (metres) squared; *cm = centimetres; Kg=kilograms; Cont.=continuous; dis./cont.= discrete treated as continuous; cat.= categorical. Note that for the Musculoskeletal Examination and Strength/Power factors, positive values indicate greater left limb values compared to right limb values; negative values indicate greater right limb*
