## SUPPLEMENTARY FILE 4 for "Can prognostic factors for indirect muscle injuries in elite football (soccer) players be identified using data from preseason screening? An exploratory analysis using routinely-collected periodic health examination records"

**Supplementary file 4: Graph to show the distribution of observed and imputed values for continuous variables**

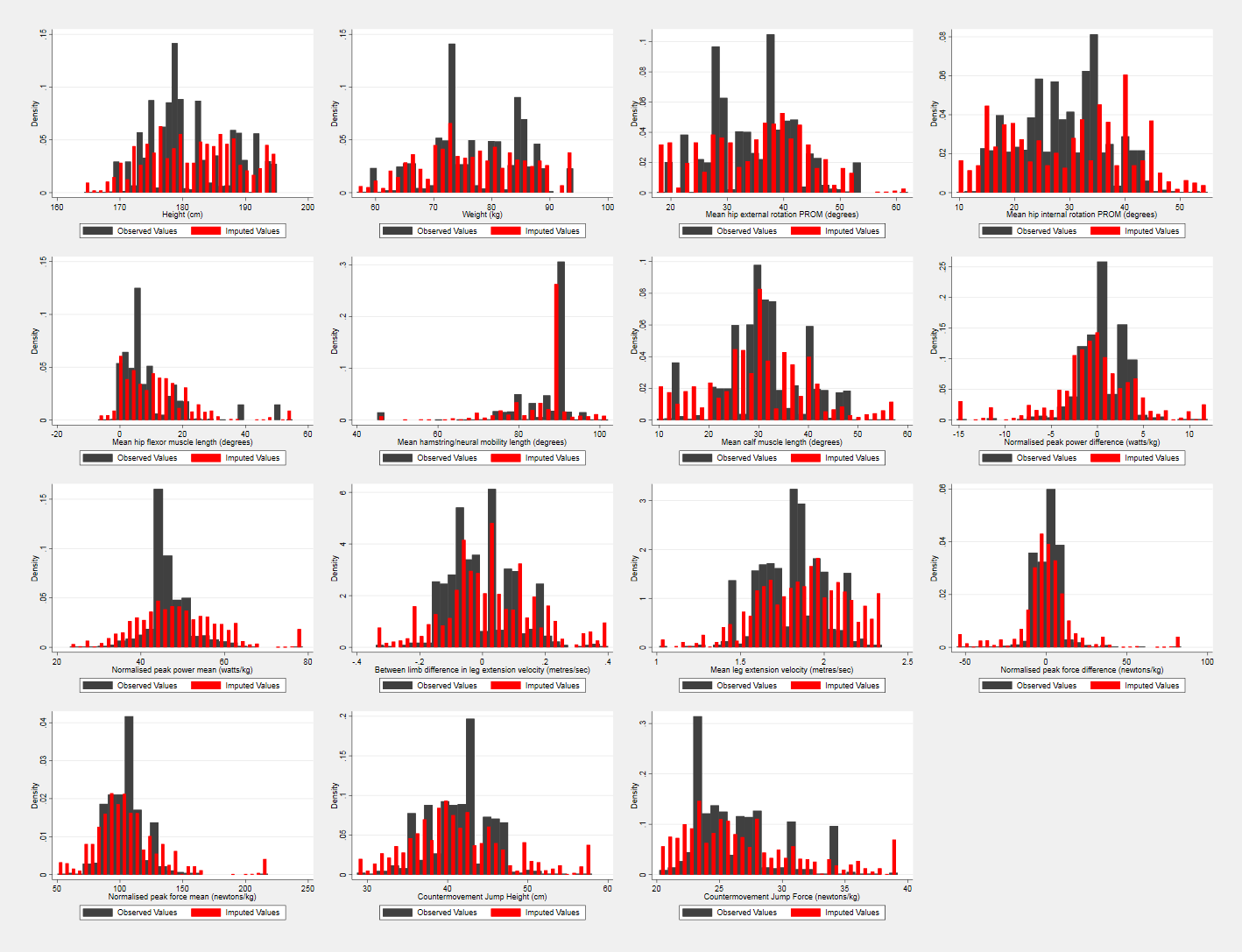

Key: cm = centimetres;

kg = kilograms;

PROM = passive range of movement
