## SUPPLEMENTARY FILE 7 for "Can prognostic factors for indirect muscle injuries in elite football (soccer) players be identified using data from preseason screening? An exploratory analysis using routinely-collected periodic health examination records"

**Supplementary file 7: Results of univariable and multivariable analyses – Sensitivity analysis using complete case data**

|  | Univariable (unadjusted) | | | | Multivariable (adjusted for age, height, weight) | | | |
| --- | --- | --- | --- | --- | --- | --- | --- | --- |
| **Candidate PF & Type** | OR | 95% CI | P | Best model fit | OR | 95% CI | P | Best model fit |
| **Anthropometric (Adjustment Factors):** |  |  |  |  |  |  |  |  |
| 1: Age (years) | **1.15** | 1.08 to 1.22 | **<0.00** | **Linear** | **1.14** | **1.64 to 1.22** | **<0.00** | **Linear** |
| 2: Height (cm) | 1.02 | 0.98 to 1.05 | 0.36 | - | 0.99 | 0.94 to 1.05 | 0.73 | - |
| 3: Weight (kg) | **1.04** | **1.01 to 1.08** | **0.01** | **Linear** | 1.01 | 0.96 to 1.06 | 0.60 | - |
| **Within 3 years preceding PHE, frequency of:** |  |  |  |  |  |  |  |  |
| 4: foot/ankle injuries | 1.03 | 0.86 to 1.25 | 0.73 | - | 1.05 | 0.85 to 1.30 | 0.63 | - |
| 5: hip/groin injuries | 1.09 | 0.83 to 1.43 | 0.54 | - | 1.23 | 0.92 to 1.65 | 0.16 | - |
| 6: knee injuries | 0.91 | 0.67 to 1.23 | 0.54 | - | 0.93 | 0.67 to 1.28 | 0.65 | - |
| 7: shoulder injuries | 1.91 | 0.80 to 4.59 | 0.15 | **-** | 1.23 | 0.46 to 3.24 | 0.69 | **-** |
| 8: lumbar spine injuries | 0.78 | 0.53 to 1.16 | 0.22 | - | 0.96 | 0.63 to 1.46 | 0.84 | - |
| 9: iliopsoas IMIs | 0.74 | 0.36 to 1.50 | 0.40 | - | 0.86 | 0.41 to 1.79 | 0.68 | - |
| 10: hip adductor IMIs | 1.46 | 0.93 to 2.30 | 0.10 | - | 1.18 | 0.71 to 1.95 | 0.53 | - |
| 11: hamstring IMIs | **2.39** | **1.60 to 3.69** | **<0.00** | **Linear** | **1.72** | **1.09 to 2.73** | **0.02** | **Linear** |
| 12: quadriceps IMIs | 1.12 | 0.67 to 1.86 | 0.66 | - | 1.05 | 0.61 to 1.82 | 0.90 | - |
| 13: calf IMIs | **2.17** | **1.13 to 4.16** | **0.02** | **Linear** | 1.52 | 0.69 to 3.33 | 0.30 | - |
| **Within 3 years preceding PHE, most recent:** |  |  |  |  |  |  |  |  |
| 14: foot/ankle injury (never) | ref | ref | ref | - | ref | ref | ref | - |
| 14: foot/ankle injury (<6 months) | 1.24 | 0.46 to 3.32 | 0.67 | - | 1.52 | 0.69 to 3.34 | 0.30 | - |
| 14: foot/ankle injury (6-12 months) | 1.65 | 0.46 to 6.00 | 0.45 | - | 1.32 | 0.54 to 3.21 | 0.54 | - |
| 14: foot/ankle injury (>12 months) | 1.90 | 0.71 to 4.97 | 0.20 | - | 1.12 | 0.58 to 2.14 | 0.74 | - |
| 15: hip/groin injury (never) | ref | ref | ref | - | ref | ref | ref | - |
| 15: hip/groin injury (<6 months) | 0.77 | 0.31 to 1.91 | 0.57 | - | 1.30 | 0.50 to 3.38 | 0.60 | - |
| 15: hip/groin injury (6-12 months) | 0.55 | 0.31 to 1.91 | 0.23 | - | 0.72 | 0.25 to 2.05 | 0.54 | - |
| 15: hip/groin injury (>12months) | 1.56 | 0.79 to 3.09 | 0.20 | - | 1.79 | 0.87 to 3.69 | 0.11 | - |
| 16: knee injury (never) | ref | ref | ref | - | ref | ref | ref | - |
| 16: knee injury (<6 months) | 0.88 | 0.31 to 2.55 | 0.82 | - | 0.92 | 0.27 to 3.12 | 0.89 | - |
| 16: knee injury (6-12 months) | 1.20 | 0.51 to 2.82 | 0.68 | - | 1.14 | 0.46 to 2.82 | 0.77 | - |
| 16: knee injury (>12months) | 0.94 | 0.51 to 1.72 | 0.85 | - | 1.10 | 0.57 to 2.12 | 0.77 | - |
| 17: shoulder injury (never) | ref | ref | ref | - | ref | ref | ref | - |
| 17: shoulder injury (<6 months) | 2.14 | 0.38 to 11.89 | 0.38 | - | 1.54 | 0.24 to 9.91 | 0.65 | - |
| 17: shoulder injury (6-12 months) | 1.07 | 0.15 to 7.71 | 0.94 | - | 0.80 | 0.10 to 6.27 | 0.83 | - |
| 17: shoulder injury (>12months) | 2.49 | 0.63 to 9.87 | 0.19 | - | 1.55 | 0.37 to 6.52 | 0.548 | - |
| 18: lumbar spine injury (never) | ref | ref | ref | - | ref | ref | ref | - |
| 18: lumbar spine injury (<6 months) | 0.93 | 0.23 to 3.80 | 0.92 | - | 2.20 | 0.46 to 10.40 | 0.32 | - |
| 18: lumbar spine injury (6-12 months) | 0.69 | 0.15 to 3.18 | 0.91 | - | 0.84 | 0.17 to 4.06 | 0.83 | - |
| 18: lumbar spine injury (>12months) | 0.55 | 0.26 to 1.14 | 0.11 | - | 0.75 | 0.34 to 1.66 | 0.48 | - |
| 19: iliopsoas IMI (never) | ref | ref | ref | - | ref | ref | ref | - |
| 19: iliopsoas IMI (<6 months) | 1.18 | 0.07 to 19.09 | 0.91 | - | 1.04 | 0.06 to 18.44 | 0.98 | - |
| 19: iliopsoas IMI (6-12 months) | 0.47 | 0.09 to 2.48 | 0.38 | - | 0.53 | 0.10 to 2.85 | 0.46 | - |
| 19: iliopsoas IMI (>12months) | 0.89 | 0.30 to 2.63 | 0.83 | - | 0.80 | 0.26 to 2.46 | 0.70 | - |
| 20: hip adductor IMI (never) | ref | ref | ref | - | ref | ref | Ref | - |
| 20: hip adductor IMI (<6 months) | 1.49 | 0.55 to 4.00 | 0.43 | - | 1.12 | 0.38 to 3.30 | 0.84 | - |
| 20: hip adductor IMI (6-12 months) | 1.59 | 0.47 to 5.36 | 0.46 | - | 1.23 | 0.32 to 4.76 | 0.69 | - |
| 20: hip adductor IMI (>12months) | 1.76 | 0.71 to 4.36 | 0.22 | - | 1.43 | 0.47 to 4.35 | 0.531 | - |
| 21: hamstring IMI (never) | ref | ref | ref | - | ref | ref | ref | - |
| 21: hamstring IMI (<6 months) | 4.06 | 0.80 to 20.63 | 0.09 | - | 1.64 | 0.27 to 9.99 | 0.59 | - |
| 21: hamstring IMI (6-12 months) | 1.76 | 0.80 to 20.63 | 0.09 | - | 1.25 | 0.50 to 3.13 | 0.64 | - |
| 21: hamstring IMI (>12months) | **4.73** | **2.05 to 10.93** | **<0.00** | **Linear** | **2.94** | **1.15 to 7.53** | **0.03** | **Linear** |
| 22: quadriceps IMI (never) | ref | ref | ref | - | ref | ref | ref | - |
| 22: quadriceps IMI (<6 months) | 4.11 | 0.45 to 37.37 | 0.21 | - | 4.03 | 0.41 to 39.22 | 0.23 | - |
| 22: quadriceps IMI (6-12 months) | 0.51 | 0.15 to 1.76 | 0.29 | - | 0.66 | 0.18 to 2.39 | 0.52 | - |
| 22: quadriceps IMI (>12months) | 1.21 | 0.52 to 2.83 | 0.65 | - | 0.89 | 0.35 to 2.28 | 0.81 | - |
| 23: calf IMI (never) | ref | ref | ref | - | ref | ref | ref | - |
| 23: calf IMI (<6 months) | 4.44 | 0.92 to 21.36 | 0.06 | **-** | 3.33 | 0.64 to 17.30 | 0.15 | - |
| 23: calf IMI (>6 months)* | 1.66 | 0.66 to 4.23 | 0.28 | - | 0.88 | 0.29 to 2.76 | 0.84 | - |
| **Musculoskeletal:** |  |  |  |  |  |  |  |  |
| 24. Mean PROM hip IR (deg.) | **0.97** | **0.94 to 0.99** | **0.01** | **Linear** | 0.98 | 0.95 to 1.00 | 0.10 | **-** |
| 25. Mean PROM hip ER (deg.) | **0.96** | **0.93 to 0.99** | **0.02** | **Linear** | 0.98 | 0.94 to 1.02 | 0.26 | - |
| 26. Mean hip flexor length (deg.) | 1.02 | 0.99 to 1.05 | 0.21 | - | 1.03 | 0.99 to 1.06 | 0.13 | - |
| 27. Mean hamstring/neural mobility length (deg.) | 0.98 | 0.95 to 1.02 | 0.40 | - | 0.96 | 0.92 to 1.00 | 0.06 | - |
| 28. Mean calf muscle length (deg.) | 0.99 | 0.96 to 1.02 | 0.38 | - | 0.98 | 0.95 to 1.01 | 0.21 | - |
| **Strength/Power:** |  |  |  |  |  |  |  |  |
| 29: Max. leg extension power difference (W/kg^-0.67^) | 0.98 | 0.90 to 1.06 | 0.63 | - | 0.97 | 0.89 to 1.06 | 0.59 | - |
| 30: Mean of max. leg extension power (W/kg^-0.67^) | 1.03 | 0.99 to 1.06 | 0.16 | - | 1.00 | 0.97 to 1.05 | 0.81 | - |
| 31: Max. leg extension velocity difference (m.s ^-1^) | 2.64 | 0.31 to 22.58 | 0.38 | - | 3.87 | 0.39 to 38.79 | 0.25 | - |
| 32: Mean of max. leg extension velocity (m.s ^-1^) | 2.49 | 0.74 to 8.32 | 0.14 | - | 1.60 | 0.42 to 6.09 | 0.49 | - |
| 33: Max leg extension force difference (N/kg^-0.67^) | 0.99 | 0.97 to 1.01 | 0.32 | - | 0.99 | 0.97 to 1.01 | 0.33 | - |
| 34: Mean of max. leg extension force (N/kg^-0.67^) | 1.00 | 0.99 to 1.01 | 0.98 | - | 1.00 | 0.98 to 1.01 | 0.59 | - |
| 35: CMJ Force per kg of body mass (N/kg) | 0.98 | 0.91 to 1.05 | 0.58 | - | 0.97 | 0.89 to 1.05 | 0.47 | - |
| 36: CMJ height (cm) | 1.03 | 0.97 to 1.09 | 0.35 | - | 1.00 | 0.95 to 1.07 | 0.86 | - |

*Key: PHE= periodic health examination; PF= prognostic factor; OR=odds ratio; CI=confidence interval; ref=reference category; I-IMI=index indirect muscle injury; IMI= indirect muscle injury; Freq= frequency; WBL=weight bearing lunge; CMJ=countermovement jump; PROM=passive range of movement; deg. = degrees; SLR= straight leg raise; BMI= body mass index;* kg/m^2^ = kilograms/body height (metres) squared; *cm = centimetres; Kg=kilograms; W= watts (note: W/kg^-0.67^ has a scaling factor to normalise force to body mass[42]); N= newtons (note: N/kg^-0.67^ has a scaling factor to normalise force to body mass);max.=maximum; m.s.= metres per second; cm = centimetres; Kg=kilograms; - = not applicable; * indicates merged category (using calf IMI (6-12 months) and calf IMI (>12months). This was required due to the occurrence of perfect predictions in the Calf (6-12 months) category as a consequence of reduced sample size.* ***Note:*** *ORs are expressed per one-unit increase for all continuous factors, and according to category for all categorical factors; Factors in* ***bold*** *indicate significance at the 0.05 level.*
